# Diffusion tractography predicts Deep Brain Stimulation evoked potential amplitude and delay

**DOI:** 10.1101/2024.04.11.24305627

**Authors:** Sumiko Abe, Jessica Vidmark, Estefania Hernandez-Martin, Maral Kasiri, Rahil Sorouhmojdehi, S. Alireza Seyyed Mousavi, Terence D. Sanger

## Abstract

**Objective:** This study investigated the relationship between DBS evoked potentials (EPs) and diffusion tensor imaging (DTI) in a group of patients with dystonia who underwent DBS treatment. EPs and DTI are both useful methods for studying neural connectivity in the brain but measure different aspects of brain function. EPs provide information on electrical connectivity, while DTI provides information on anatomical pathways connecting regions.

**Methods:** This study focused on the pallidum and motor thalamus nuclei, which are common targets for DBS in dystonia. Prior to DBS implantation, DTI images were acquired for each patient, and were processed to obtain DTI coefficients such as length (L), volume (V), and fractional anisotropy (FA) of the fiber tracts. The relationship between the fiber tracts and electrophysiology was examined using a generalized linear model (GLM).

**Results:** We showed that the amplitude of EPs correlated with FA and tract volume, while delay correlated with tract length. These findings suggest that DBS signals travel across tracts to affect both local and distant brain regions, and the magnitude of the effect of DBS is determined by the integrity of the white matter tract, while DBS signal delay is affected by the tract length. Our results further suggest that the magnitude and delay of the spread of the DBS signal may be predicted by the DTI connectivity. This provides strong supporting evidence for other studies that have assumed, but have been unable to test, such a relationship.

**Conclusion:** Overall, this study suggests that the electrical effects of DBS can be at least partially predicted by noninvasive DTI imaging in patients with dystonia. By combining EPs with DTI, we could investigate the propagation of stimulation pulses through brain regions. While this relationship has been previously hypothesized by the neuroscience community, this is the first study in humans to demonstrate this relationship between DBS EPs and DTI, thereby advancing the field of human brain mapping and enhancing the precision of neurosurgical targeting.

## I. Introduction

Deep Brain Stimulation (DBS) is a neurosurgical procedure used to treat movement disorders such as dystonia [1], which is characterized by involuntary muscle contractions that lead to repetitive movements or abnormal postures [2]–[4]. DBS is effective in improving dystonic symptoms by modulating abnormal neural activity in the pallidum and thalamus, two brain regions that are commonly targeted during this procedure, for children with dystonia [5]–[8].

Recent clinical development involves the utilization of temporary stereoelectroencephalography (sEEG) leads for determining optimal DBS targets for dystonia [9], [10]. This procedure allows us to record intracranial signals from deep brain regions during stimulation and while the patient is awake. Using this approach allows us to record DBS evoked potentials (EPs), which provides us with the opportunity to gain insights into how electrical stimulation modulates neural activity in distant regions [11]. An important challenge in the quantification of intracranial EPs is the short delay (often less than 1 ms), often causing the EP to be obscured by the stimulation artifact. Thus, utilizing proper tools for quantifying intracranial EPs is essential.

Diffusion tensor imaging (DTI) is a technology that studies white matter structures in the brain. It measures the diffusion of water molecules and provides information about the direction and integrity of white matter tracts [12]. DTI is particularly valuable in assessing anatomical connectivity between different brain regions, providing details on the morphology of the tracts. Despite the extensive research on DTI technique and its applications, including optimization of DBS target and improvement of surgical outcomes [13]–[16], the relationship between DTI parameters and the propagation of DBS signals has never been confirmed.

Therefore, we herein aim to study the relationship between DTI parameters and DBS signal propagation in order to improve our understanding of DBS impact on neural activity [17]. This paper presents findings from a comprehensive study into the correlation between DTI parameters and DBS EP characteristics - specifically, the EP amplitude and delay. In this study, we analyzed DBS EPs and DTI measures from 12 children with dystonia undergoing DBS procedure. First, we outline patient selection and the surgical procedure. Second, we discuss the details of the electrophysiological recording process, the stimulation protocol, and the steps taken in the processing of electrophysiological data. Third, we explain the image processing, the electrode trajectory construction, and the tractography process. Finally, we present our statistical analyses and results, and discuss our findings.

## II. Materials and Methods

### A. Subjects

Twelve (12) adolescent patients undergoing DBS surgery for the treatment of dystonia participated in this study (Table I). The diagnosis of dystonia was established by a pediatric movement disorder specialist (T.D.S.) using standard criteria [18]. All patients provided signed informed consent for surgical procedures in accordance with standard hospital practice at Children’s Health Orange County (CHOC) or Children’s Hospital Los Angeles (CHLA). The study protocol was approved by the institutional review boards (IRB) of both hospitals. The patients, or parents of minor patients, also signed an informed consent to the research use of electrophysiological data and provided Health Insurance Portability and Accountability Act (HIPAA) authorization for the research use of protected health information.

**TABLE I:**
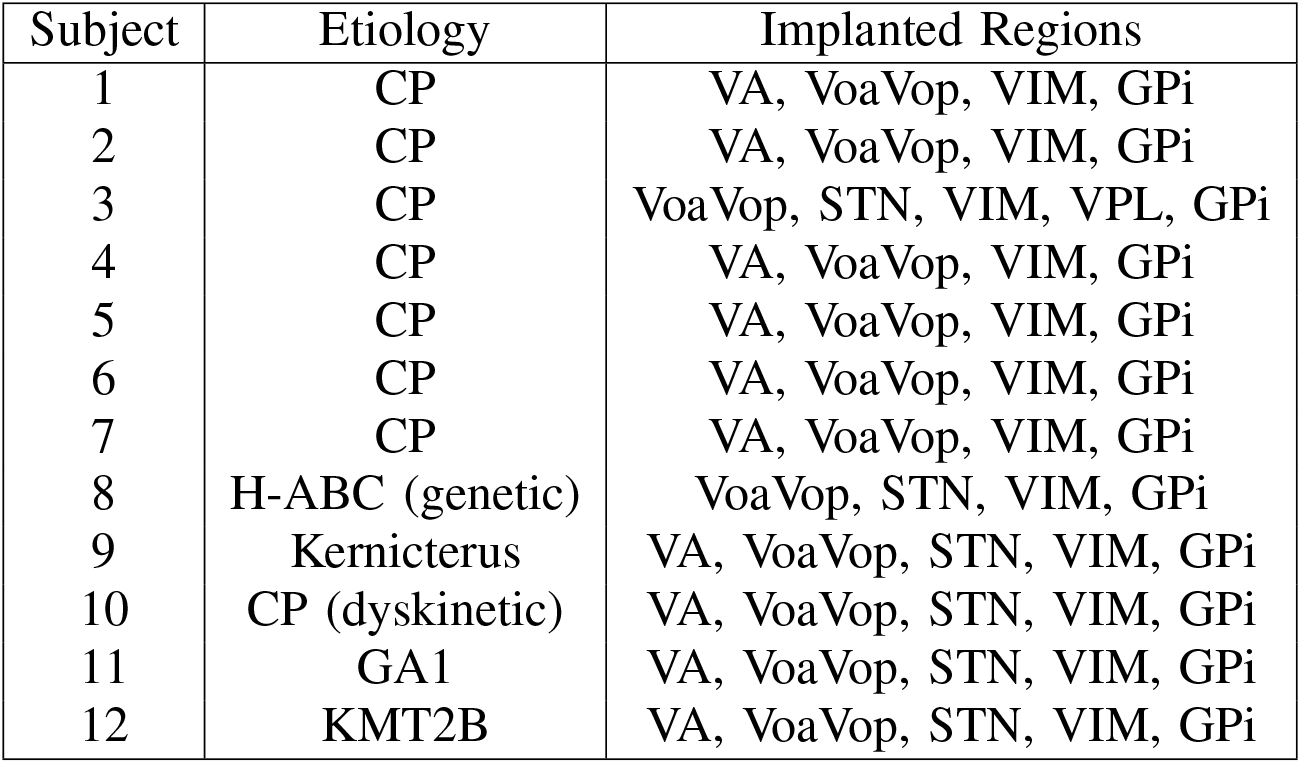
Demographic characteristics, 5 female and 7 male patients, age between 10-20 years old. CP: Cerebral palsy, H-ABC: Hypomyelination with atrophy of the basal ganglia and cerebellum, GA1: Glutaric aciduria type 1, GPi: globus pallidus internus; VoaVop: ventral oralis anterior-posterior; STN: subthalamic nucleus; VA: ventral anterior; VPL: ventral posterolateral, VIM: ventral intermediate; PPN: pedunculopontine nucleus.

### B. Surgical Procedure and Depth Recordings

We implanted up to 10 temporary Adtech MM16C depth leads (Adtech Medical Instrument Corp., Oak Creek, WI, USA) at potential DBS targets, as part of our clinical procedure for determining DBS targets. Based on prior studies of clinical efficacy in children with dystonia, typical targets for DBS include the subthalamic nucleus (STN) and globus pallidus internus (GPi) in the basal ganglia, as well as the ventral intermediate (VIM), ventral oralis anterior-posterior (VoaVop), ventral anterior (VA), and ventral posterolateral (VPL) thalamic subnuclei [10], [19]. Depth electrodes were placed using standard stereotactic procedures, with the most distal stimulation contact placed at the target location [20]. Electrode location was confirmed through co-registration of the preoperative magnetic resonance imaging (MRI) scan with the postoperative computed tomography (CT) scan. Thalamic targeting was further confirmed through identification of leads in subnuclei known to have greater or lesser response to median nerve electrical stimulation [21].

### C. Electrophysiological Recordings

Recordings were performed during the first 24 to 48 hours after clinical implantation of the temporary sEEG depth leads. Each sEEG lead has a diameter of 1.2 mm and contains 6 low-impedance (1–2 *k*Ω) ring macro-contacts with 2 mm height and 5 mm on-center spacing, as well as 10 high-impedance (70*−*90 *k*Ω) micro-contacts (50 *µm* diameter). The micro-contacts are arranged in groups of 2 or 3, spaced evenly around the circumference of the electrode shaft, halfway between neighboring pairs of ring macro-contacts. The external proximal ends of the leads were connected to Adtech Cabrio™ connectors modified to include a custom unity-gain preamplifier for each microwire electrode to reduce noise and motion artifacts. Macro-contacts bypass the preamplifiers to allow for external electrical stimulation. All data reported here are from the high impedance micro-contact recordings, while stimulating through low-impedance macro-contacts. All signals were amplified, sampled at 24 kHz, and digitized by a Tucker-Davis Technologies PZ5M analog-to-digital amplifier connected to an RZ2 digital signal processor. Data were streamed and stored to an RS4 high-speed data storage unit, controlled by Synapse software (System3, Tucker-Davis Technologies Inc., Alachua, FL, USA).

#### 1) Stimulation protocol

Approximately 1200 pulses with 90-*µs* width, 3 V amplitude at 25 Hz were administered through two adjacent macro-contacts (anode and cathode). In order to reduce stimulation artifacts, stimulation was performed twice for each contact pair, with the cathode and anode exchanged. When the resulting signals are added, the opposite artifact polarities cancel out while the evoked response (which does not change polarity) is augmented [22].

#### 2) Electrophysiology analysis

All data processing was performed in MATLAB2020a (The MathWorks, Inc., Natick, MA, USA). The recorded neural activity was defined as the relative response between two nearby micro-contacts (i.e., local bipolar recordings) and was time-averaged and time-locked to a stimulus artifact. To ensure accuracy and avoid time delays in synchronization pulses between different equipment, the stimulus time was identified by looking for the occurrence of stimulus artifacts in the recorded data. We upsampled the raw data to 120 kHz, detected the stimulus artifacts and split them into 11-ms segments. All stimulus artifacts in all segments were then aligned to each other at time “0” using cross-correlation and averaged. This stimulus-triggered averaging procedure hence greatly increased the signal-to-noise ratio (SNR) of the final average neural response. The above process was repeated for both stimulation polarities. In order to detect the evoked potentials (EPs), we employed a novel algorithm for automated detection of polarity reversed EPs (ADPREP) [22]. The algorithm starts by identifying and removing decay artifacts following the main stimulus artifact. This is achieved by fitting a sum of two exponential decays to each of the two stimulation types [23]. The resulting cathodic and anodic recordings were averaged to obtain the artifact-reduced neural response (Fig.1). Subsequently, the algorithm determines whether a significantly positive correlation exists between the artifact-reduced cathodic and anodic recordings. Furthermore, it assesses the SNR of the resulting average response in this correlated region, using an empirically chosen threshold. Only recordings meeting these criteria are labeled as EPs and included in the analysis of this study. Detected EPs were characterized by their peak-to-peak amplitude (P2P) and time-to-(first-) peak delay (T2P) as shown in Fig.1 and grouped within each micro-contact row for further analysis.

**Fig. 1:**
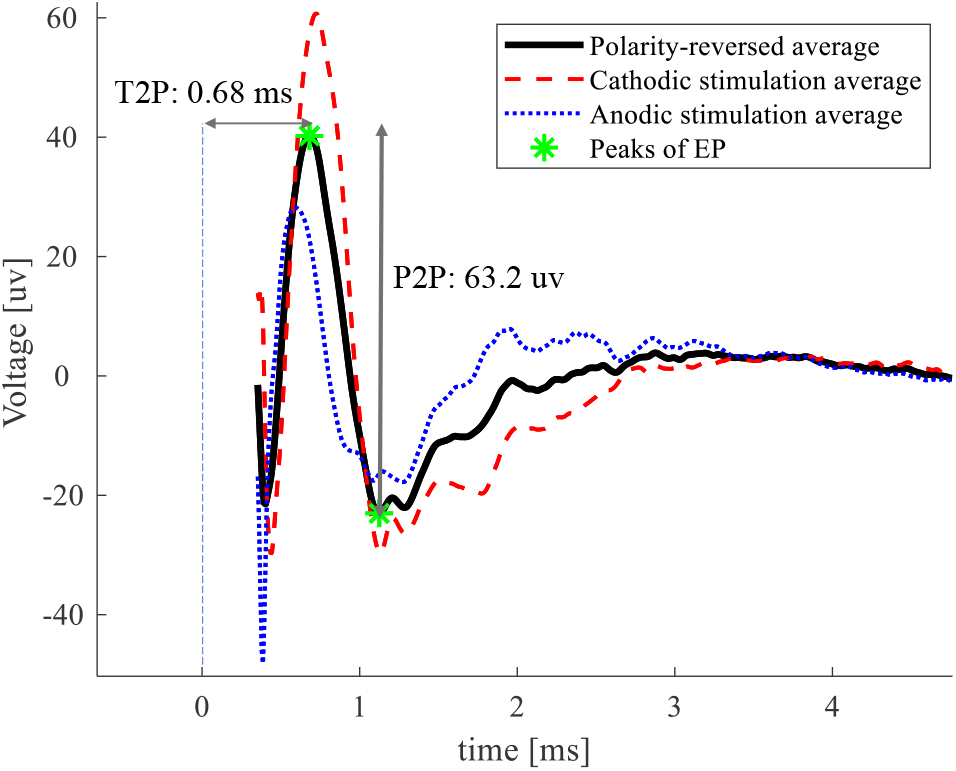
Illustration of EP detected from the polarity-reversed average VoaVop recording during GPi stimulation. Employing the ADPREP algorithm, the decay artifacts was removed from cathodic (red dashed line) and anodic (blue dashed line) recordings and resulting artifact-reduced signals were averaged to generate the final neural response (black line). P2P amplitude and T2P delay of the detected EP are depicted here.

### D. Imaging

#### 1) Image Data Acquisition

Preoperative T1-weighted (structural MRI) volumes were obtained using a MAGNETOM 3T (SIEMENS Medical System, Erlangen, Germany) MRI scanner for precise anatomical localization. The acquisition settings included a repetition time (TR) of 1800 ms, echo time (TE) of 2.25 ms, flip angle of 8^*°*^, voxel size of 1mm^3^, and a field of view (FOV) of 240mm^2^. In addition, preoperative diffusion-weighted imaging (DWI) sequences suitable for tractography were acquired.

Two DWI (diffusion tensor imaging) scans with opposite phase-encoded polarity were obtained, along with calibration scans, to correct for magnetic inhomogeneities [24], which could otherwise lead to errors in tractography reconstruction. Consequently, two sequences were acquired, one with an anterior-posterior direction and the other with a posterior-anterior direction, to address the phase encoding artifact in EPI (echo plannar imaging) sequence. The parameters for these sequences were as follows: flip angle of 90^*°*^, TR of 9000 ms, TE of 113 ms, voxel size of 2.5 × 2.5 × 2.5 mm^3^, FOV of 250 × 250 mm^2^, b-value of 1000 s*/*mm^2^, and 36 gradient directions. CT volumes were obtained using a GE (GENERAL ELECTRIC Healthcare, Milwaukee, WI, USA) scanner with a size of 512 × 512 × 320mm^3^ to confirm the localization of the sEEG leads.

#### 2) Image Processing

Diffusion images were processed using the TOP-UP algorithm [25] and Eddy-current process [26] to correct distortions and motion artifacts. Then, both post-surgery CT and pre-surgery DTI volumes were aligned to the same plane of T1 images which has been resliced to anterior commissure-posterior commissure line (ACPC line) using the FLIRT tool in FSL [27]. The voxel size of the acquired images were 1mm^3^.

Studies on healthy, adult images use tools like ANTS [28] and FSL [29] to warp the brain; however, in our study population, the brains are asymmetric, distorted with smaller dimensions.

In order to precisely segment all the components (subregions) of thalamus and pallidum which encompasses our sEEG targeting, first, we used Freesurfer software [30] to segment the brain images to localize pallidum and thalamus for each patient. However, this segmentation is not perfect and needs manual adjustment using DSI-studio [31]. Therefore, we calculated a transformation matrix for each parent region (pallidum and thalamus) from Montreal Neurological Institute (MNI) space to individual space by FSL FLIRT tool. Next, we applied the same transformation matrix on the sub-regions from DISTAL atlas (built by LEAD-DBS [32]), as it contains all components of pallidum and thalamus and matches with MNI template for all these components (other atlases usually do not).

These procedures allowed us to visualize the segmented sub-regions, the leads which were reconstructed by post-surgery CT scan, pre-surgery DTI volumes and MRI T1 anatomic volume in each individual space using the DSI Studio visualization package. We integrated these images to perform DTI tractography between origin and target contacts.

#### 3) sEEG lead trajectory reconstruction

Standard lead trajectory software [33] cannot be utilized in our case since our sEEG lead electrode specifications do not conform with established standards such as Boston Scientific Inc. or Medtronic Inc. leads. Moreover, standard software do not support visualizing of 10 to 12 leads simultaneously. Therefore, after co-registering the CT images to the structural MRI volume (T1-weighted), we calculated projection images [34] to accurately place the sEEG leads. In other words, we projected 3D voxel values onto a 2D image, capturing the prominent attenuation value (Fig.2, top), following the equation:

**Fig. 2:**
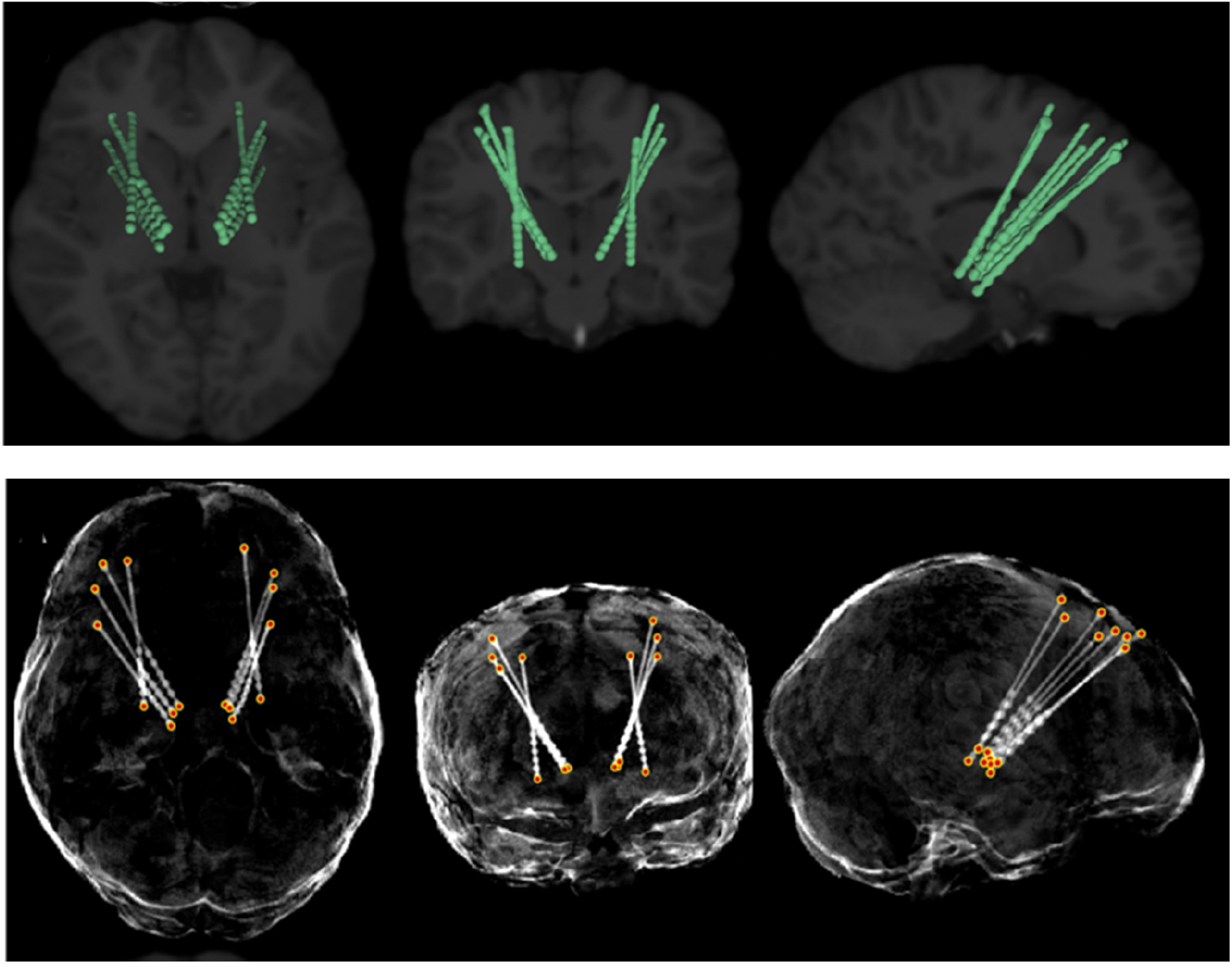
sEEG lead trajectory reconstruction in axial (left), coronal (middle), and sagittal (right) views in an individual space. (Top) The CT scan is marked as a dense structure and fitted onto the structural images. (Bottom) Projection images in axial, coronal, and sagittal views are used to reconstruct the positions of the sEEG lead electrodes.

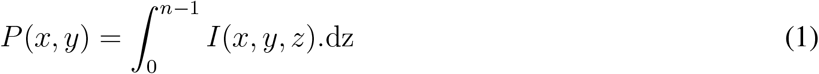

where n is the number of slices, *x* and *y* refer to coordinates of the voxel in one slice and *z* is the coordinate in the slice axis. Using this method, each X-Y (axial) coordinate represents only the pixel with the prominent value along the Z-axis. The same can be obtained on the Y-Z (sagittal) and Z-X (coronal) planes. This allows us to observe all dense structures within each volumetric area in a single 2D image (Fig.2, top), also enabled us to obtain precise spatial coordinates of multiple sEEG leads. Using DSI-studio package the sEEG leads coordinates were integrated with atlas images as shown in Fig.3.

**Fig. 3:**
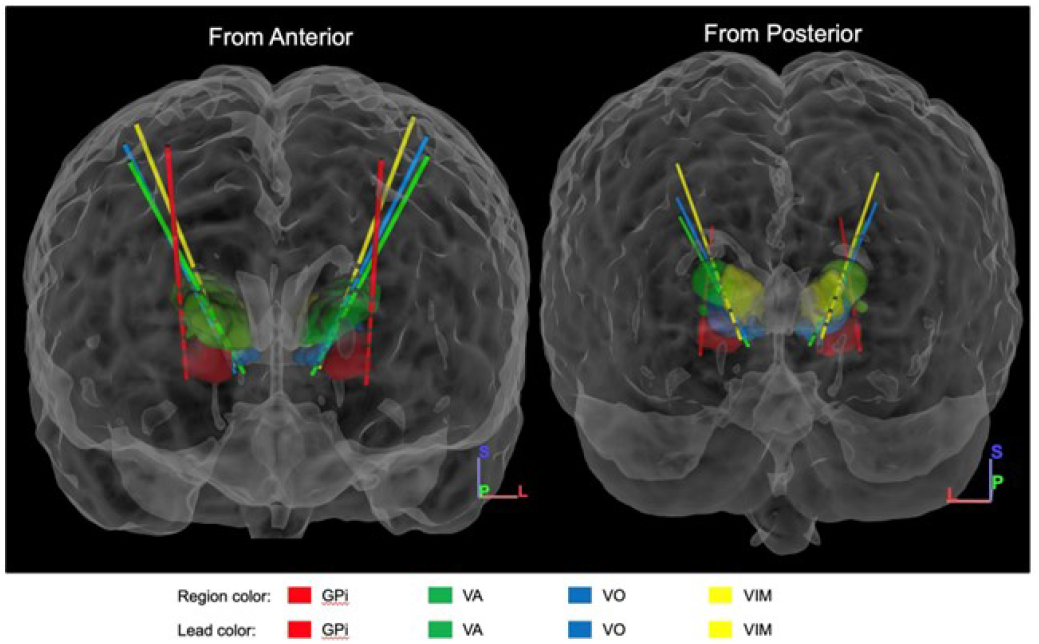
The segmented DBS targets including GPi (red), VA (green), VoaVop (blue), and VIM (yellow) nuclei along with the reconstructed sEEG leads trajectories are depicted using DSI-Studio package.

#### 4) Tractography Seed and Target Selection

We utilized the metal artifacts of the macro-contacts on Adtech leads (Fig.4, middle) to estimate the tractography seeds. The post-surgery CT scans were thresholded to detect metal artifacts by a traditional approach [35], enabling the generation of a binary mask that provided the spatial localization of the macro-contacts for all sEEG leads.

**Fig. 4:**
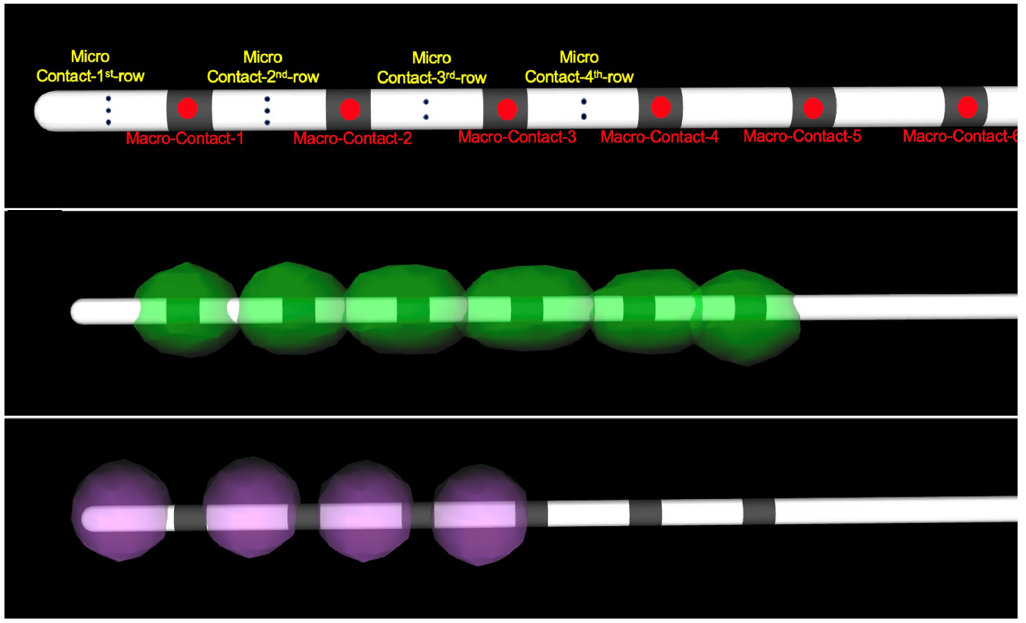
sEEG lead electrodes with sample micro- and macro-contacts. (Top) The Adtech MM16C sEEG lead. (Middle) Macro-contacts, detected from CT metal artifacts, are depicted as green spheres. (Bottom) Violet spheres represent the reconstructed region for the micro-contact rows based on the linear interpolation.

In addition to identifying the macro-contacts locations, we also identified the micro-contacts locations through which all the EPs were recorded. The center coordinates of micro-contacts between macro-contacts were calculated by a linear interpolation based on the vendor product specification. Then, the micro-contacts were represented as spheres area, with 3 mm diameter, surrounding the centers (Fig.4, bottom, in purple color).

#### 5) Fiber Tracking

DTI was employed to reconstruct the tractography, which is a technique commonly utilized to investigate fiber orientations and quantify diffusion properties [36]. In our study, we applied q-space diffeomorphic reconstruction [37] for DTI fiber tracking using DSI-Studio. The determination of the fiber tracking threshold was based on quantitative anisotropy (QA) coefficients [36]. Within each macro-contact region (Fig.2, middle), we randomly generated seeds with an average of 1000 seeds per voxel and a turning angle of 10^*°*^.

The macro-contact metal artifacts from the CT scans [35] were visualized in DSI-Studio and fused with the deep brain atlas (DISTAL atlas). In order to select the seeds, we conducted a conjunction analysis between the deep brain atlas and reconstructed macro contact areas . In this way, the macro-contact regions located within each nucleus were identified. Fiber tracking was conducted to visualize the neural pathway connecting the GPi to the VoaVop region, allowing us to reconstruct the corresponding white matter fiber tracts. Starting from the GPi region, the fiber tracts were traced and followed until they reached the VoaVop region, enabling us to visualize the anatomical connection between these two regions and better understand the underlying anatomic structural connectivity. The fiber tracking results also yielded valuable information regarding the anatomical characteristics of the fiber tracts connecting the GPi and VoaVop regions, including fractional anisotropy (FA), tract length, and tract diameter [38].

### E. Statistical analysis

The DTI coefficients, such as tract length, tract diameter, and fractional anisotropy (FA), can be used to quantify the characteristics of each fiber tract. On the other hand, the EPs as a measure of DBS signal propagation can be characterized by their peak-to-peak amplitude (P2P) and time-to-(first-) peak delay (T2P). In this study, our goal was to explore the relationship between these EP characteristics and DTI coefficients. The hypothesis is that the fiber length will be positively correlated with the delay (T2P) while fiber diameter will be negatively correlated with delay and positively correlated with the amplitude (P2P). Furthermore, we hypothesize that the FA will be positively correlated with P2P and negatively correlated with T2P. In order to test these hypotheses at both individual and group level we used linear mixed effect model (LME) and generalized linear model (GLM), respectively. In our study, the GLM is defined as:

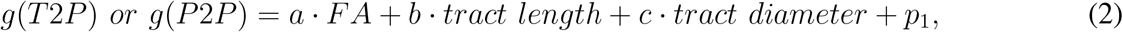

where a, b, and c are the coefficients of predictors and *p*_1_ is the intercept of the fitted line to the population data, and *g*(.) is a Gaussian link function. Moreover, we tested our hypotheses at individual level by performing a linear mixed effect model. These linear models were defined as follows:

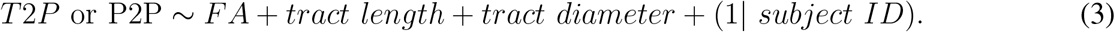

The selection of the LME model was based on a comparison of different models with varying complexities (including or excluding interactions) using analysis of variance (ANOVA test). The model with the significantly lowest Akaike Information Criterion (AIC) was chosen. In cases where the difference was not significant, the simpler model was preferred to avoid overfitting. In both selected models, subject IDs were included as random effects. This allowed for the assignment of separate regression lines to each subject, considering individual variations. The DTI parameters (FA, tract length and tract diameter) served as fixed effects in the model, contributing to the overall variance in the outcome measures; T2P or P2P. All the statistical analyses were performed in R studio using the lme4 [39], [40] and emmeans [41] packages.

## III. Results

We performed DTI fiber tracking between all sEEG lead target regions with macro-contacts as seed and micro-contacts as target for all patients. Moreover, we detected EPs recorded from micro-contact rows while macro-contacts administered stimulation, then we estimated P2P and T2P values for all these recordings. As an example, in Fig. 5 the EPs recorded from different rows of VoaVop lead when stimulating GPi macro-contacts 1&2 are depicted for one subject, along with fiber tracts between GPi macro-contacts 1&2 and different rows of VoaVop lead. Both leads are on the right hemisphere. Moreover, the relation between characteristics of EPs, P2P and T2P, and DTI coefficients, FA, tract length and tract diameter are showcased in Fig. 5.

**Fig. 5:**
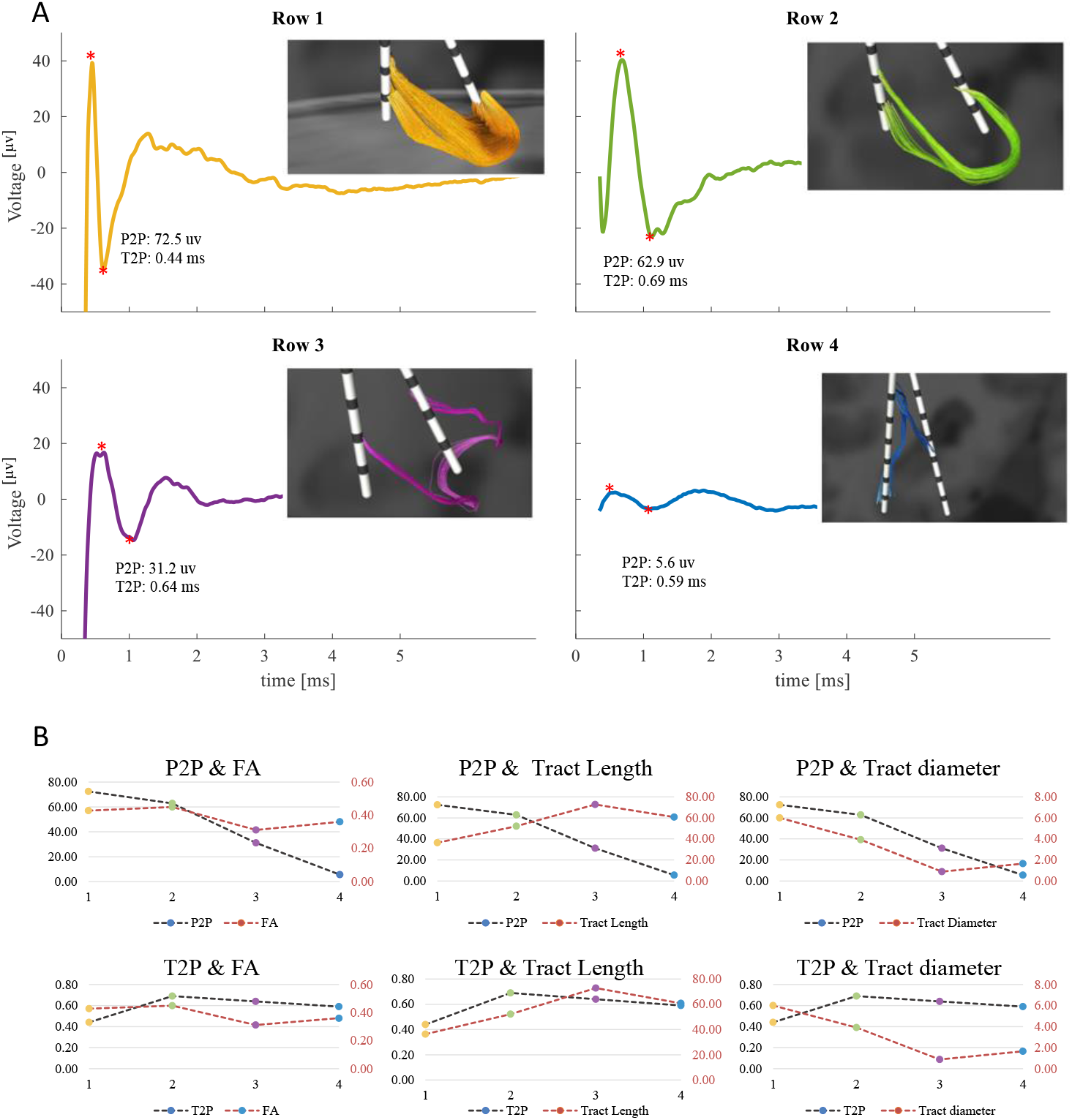
Example from the DTI-EP correlation analysis in one subject. (A) Neural recordings from micro-contacts in VoaVop (right hemisphere) during 25-Hz GPi DBS (first and second macro-contacts), along with associated fiber tracking between macro-contacts in GPi (stimulation) and micro-contact rows in VoaVop (recording) in one patient. Note how the larger the P2P amplitude the more prominent the fiber tracking. (B) Trends of EP characteristics (T2P and P2P) and DTI coefficients (length and volume of fiber tracts and FA) in the four different connections between macro-contacts in GPi to micro-contacts in VoaVop. The top row shows the relationship between P2P and DTI coefficients, whereas the bottom row shows the relationship between T2P and DTI coefficients.

All DTI coefficients, P2P, and T2P values are normalized using a min-max scaler to ensure the reliability of the models estimations. The results of the group analysis (n=12) using GLM to examine the relationship between DTI coefficients and EP characteristics (P2P and T2P) are summarized in Table II. For P2P, we observed a significant positive relationship with FA and tract diameter, indicating that higher FA and larger tract diameter were associated with larger EP amplitudes. On the other hand, there was a significant negative relationship between P2P and tract length, suggesting that longer tracts were associated with smaller P2P amplitudes (Fig. 6). In contrast, for T2P, we only found a significant negative relationship with FA, but not with the tract length and diameter. This means that higher FAs are associated with shorter EP delays (as shown in Fig. 7). At group level, these findings indicate specific associations between the DTI measures (FA, tract length and diameter) and the amplitude (P2P) and delay (T2P) of the evoked potentials.

**TABLE II:** details of GLM of EP amplitude and delay (P2P and T2P) vs. DTI coefficients; Estimation of slope, standard error, tStat, and p-value are reported for FA (a), tract length (b) and tract volume (c) and the intercept(p_1_), for both EP amplitude (P2P) and delay (T2P). Distribution: normal. 2115 observations; 2111 error degrees of freedom. Estimated Dispersion: 0.02 and 0.03 for P2P and T2P, respectively. F-statistic vs. constant model (p ≤ 0.01 for both models)

|  | P2P |  |  |  | T2P |  |  |  |
| --- | --- | --- | --- | --- | --- | --- | --- | --- |
|  | Estimate | SE | tStat | p-value | Estimate | SE | tStat | p-value |
| a | 0.12 | 0.01 | 8.76 | $\leq \mathbf{0.01}$ | -0.07 | 0.02 | -3.89 | $\leq \mathbf{0.01}$ |
| b | -0.05 | 0.01 | -3.43 | $\leq \mathbf{0.01}$ | -0.03 | 0.02 | -1.50 | 0.5 |
| c | 0.08 | 0.02 | 4.84 | $\leq \mathbf{0.01}$ | -0.03 | 0.02 | -1.38 | 0.6 |
| $p_1$ | 0.006 | 0.01 | 0.66 | 0.5 | 0.21 | 0.01 | 15.28 | $\leq \mathbf{0.01}$ |

**Fig. 6:**
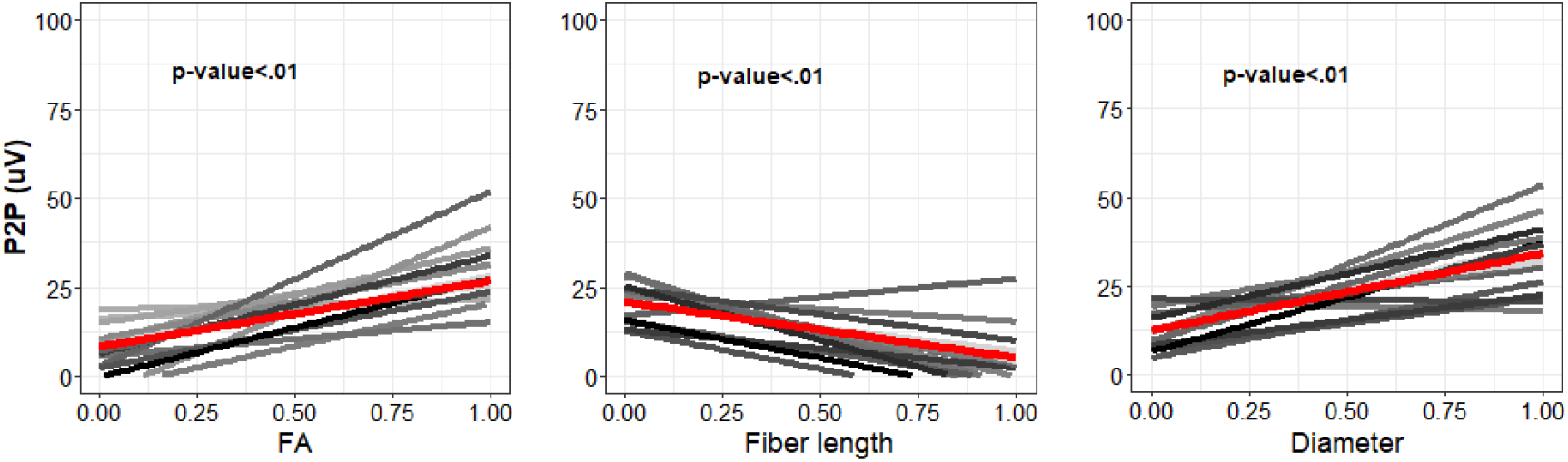
The linear relationship between DTI measures and P2P (EP amplitudes) for all subjects, represented by the LME (gray lines; each line represents one subject) and the GLM (red line) models. From left to right, the plots show the relationships of P2P with FA (*p <* 0.01), P2P with tract length (*p <* 0.01) and P2P with tract diameter (*p <* 0.01). The vertical axes represent P2P values.

**Fig. 7:**
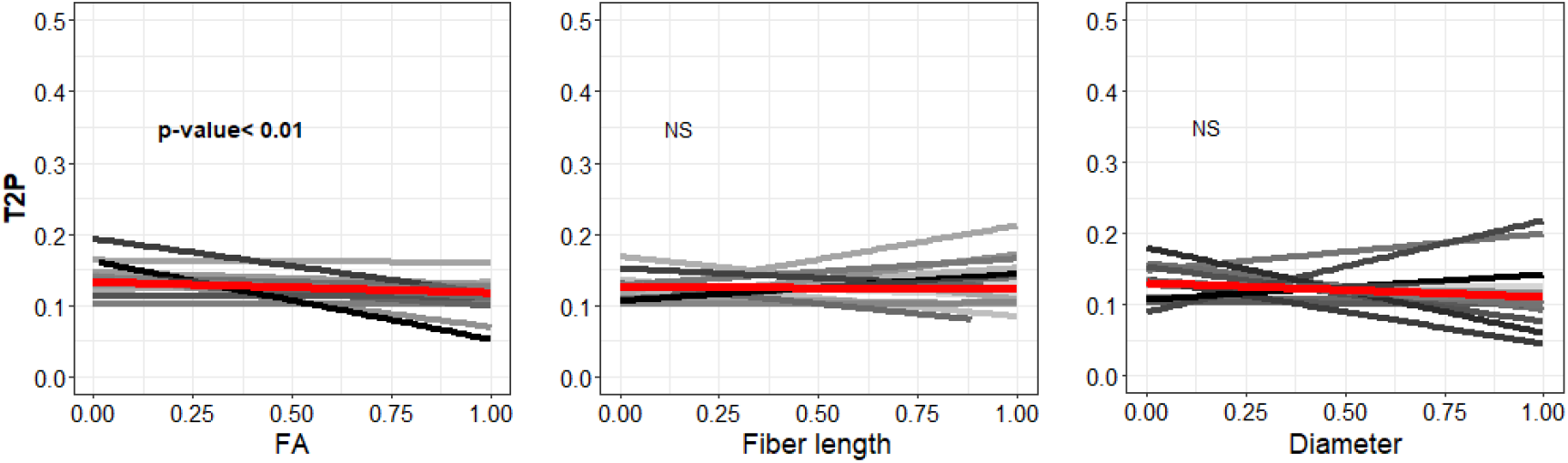
Linear mixed effect model for the relationship of DTI measure and T2P (EP delay) in the GPi-to-VoaVop neural pathway all subjects.

We performed the analysis by fitting a linear mixed effect model constructed as in equation (3) with the subject IDs as random effects and the DTI measures (FA, tract length, and diameter) as the fixed effects, and T2P or P2P as the dependent variables (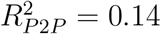 and 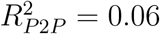), on 2115 observations (n=12).

We, then, applied type III ANOVA test with Satterthwaite’s method which revealed a significant positive correlation of P2P measure with FA (estimate= 50.56, CI [38.55 - 62.57], p-value *<* 0.01), a significant negative correlation with the tract length (estimate = -24.06, 95% CI [-37.87 – -10.25], p-value *<* 0.01), and a positive correlation with the tract diameter (estimate = 36.23, 95% CI [20.95 – 51.51, p-value *<* 0.01).

Same analysis applied to the T2P values, consisting of 2115 observations from 12 subjects (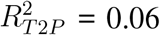 revealed only a significant negative correlation of FA with the EP T2P delays, (estimate= -0.63, 95% CI [-0.87 - -0.39], p-value *<* 0.01), and no significant correlation with the tract length (estimate= -0.10, 95% CI [-0.38 - 0.18]) or diameter (estimate= -0.24, 95% CI [-0.55 - 0.07]). All the reported p-values are corrected with Bonferroni method.

## IV. Discussion

Deep brain stimulation (DBS) has emerged as a favorable procedure in addressing treatments of several neurological disorders. It has not yet been fully understood that how DBS signal flows and propagates through white matter tracts and whether noninvasive techniques such as DTI can help with better understanding of signal propagation and characteristics of DBS evoked responses at distant brain regions. Therefore, here in this study, we investigated the relationship between DBS EPs and DTI parameters in pediatric patients and young adults with dystonia who underwent DBS treatment. Specifically, we focused on understanding how anatomical connectivity, assessed by DTI parameters, correlates with the characteristics of DBS EPs. Results from this study help us to gain insight on relationship between white matter anatomic structure (measured by DTI) and characteristics of DBS propagation (measured by EP). The observed significant positive correlation between P2P amplitude and FA and tract diameter demonstrates the relationship between the structural characteristic of white matter tracts and the efficiency or strength of neural responses induced by DBS. Furthermore, the negative correlation between P2P amplitude and tract length shows an interesting aspect that longer tracts were associated with smaller EP amplitudes. This could imply that the effectiveness of DBS-induced neural responses might be influenced by the length of transmission along these tracts. On the other hand, the significant negative relationship between T2P delay and FA shows that higher FA values correspond to shorter EP delays. This result suggests that greater white matter integrity may contribute to faster transmission or more efficient propagation of DBS-induced signals across neural pathways. However, the absence of significant correlation between T2P and tract length or diameter needs to be further explored.

Notably, these findings have several clinical implications, which potentially inform us about possible ways to optimize the DBS procedure. Understanding how DTI parameters relate to DBS EPs characteristics, ultimately results in improvement of noninvasive techniques for target selection, which leads to enhanced treatment efficacy for children with dystonia.

This study validates the physiological relevance of DTI measures by showing that anatomic connectivity does in fact predict electrical connectivity. This is not only important as a validation of advanced anatomic imaging, but it also suggests that noninvasive imaging can be used as a valid predictor of the propagation of the DBS signal from the site of stimulation to distant sites. Knowledge of this propagation may be helpful for more precise targeting of DBS electrodes when specific patterns of activation are desired.

The contributions of this investigation demonstrate further potentials in studying additional variables, such as specific neural circuitry or patient-specific characteristics, that could deepen our understanding of the observed correlations. Further study is required to assess relationship between the anatomical connections (derived from DTI measures) and intrinsic signal transmission.

## Data Availability

All data produced in the present study are available upon reasonable request to the authors

## V. Acknowledgements.

We thank our volunteers and their parents for participating in this study. We also thank Jennifer Maclean and Diana Ferman for their assistance with neurological examinations, as well as Jaya Nataraj for helping in data collection.

## VII. Authors Contributions

S.A., J.V., E.H.M., M.K., R.S., S.A.S.M., and T.D.S. conducted the experiments and collected the data. J.V. analyzed the neurophysiological recordings. S.A. performed the DTI analysis and electrode localization. R.S. integrated the neurophysiological and imaging data. M.K. performed the statistical analysis. T.D.S. performed the neurological examination. All authors contributed to writing and revision of the final manuscript.

